# Liquid Biopsy of HPV Cell-Free DNA Enables Blood-Based Early Detection and Molecular Stratification of HPV-Associated Cancer and Precancer Stages

**DOI:** 10.64898/2026.05.11.26352922

**Authors:** Qin Wang, Samuli Eldfors, Sangmi Sandra Lee, Dipon Das, Yana Al-Inaya, Gjystina Lumaj, Eliana T. Epstein, Shriya Shukla, Emma Ricart, Harsharan Dhillon, Juniper Lake, Shun Hirayama, Viktor A. Adalsteinsson, Michael G. Drage, Doga C. Gulhan, Benjamin T. Davis, Daniel L. Faden

## Abstract

Liquid biopsies targeting circulating tumor DNA enable noninvasive cancer detection but lack sensitivity in pre- and early-cancer stages, where clinical benefits would be greatest. Human papillomavirus (HPV) causes six cancer types, accounting for 5% of all cancers worldwide. Targeting HPV cell-free (cf)DNA offers a compelling opportunity to overcome current liquid biopsy constraints due to its unique tumor-specific origin, lack of sequence homology to the human genome, and the high viral-to-human copy ratio per cell. Utilizing HPV-associated anal cancer and precancer as a model, here we applied a custom, multi-feature HPV whole-genome liquid biopsy to biobanked and prospective screening cohorts spanning the HPV infection-precancer-cancer continuum. HPV cfDNA was detected years before cancer diagnosis and as early as the infection stage, with increasing detection as stages advanced. Genomic hallmarks of HPV malignancy, including HPV integration, *PIK3CA* mutations, and 3q amplification, were detected exclusively in cancer, while precancers exhibited distinct HPV genotypes. Fragmentomics analysis of HPV cfDNA revealed stage-informative signatures reflecting viral epigenetic changes during carcinogenesis. A unified classifier incorporating genomic and fragmentomics features achieved a mean AUC of 0.77 for identifying cancer and high-grade precancer, stages requiring clinical intervention. Together, these findings demonstrate the feasibility of blood-based screening and molecular risk stratification for HPV-associated cancer and precancer.

**Teaser:** Profiling blood HPV cell-free DNA detects cancer years early and distinguishes precancers needing intervention from surveillance

## INTRODUCTION

Cancers, when diagnosed and intervened early, yield markedly superior outcomes, particularly if identified during precancerous stages (*1*). However, ∼50% of cancers are still detected at advanced stages, largely due to the absence of early clinical symptoms (*1*). Liquid biopsy targeting circulating tumor DNA (ctDNA) in blood offers an attractive option for early cancer detection (*1*, *2*). Multiple biomarker modalities have been investigated in liquid biopsy, including genetic variants such as point mutations and copy number alterations (CNAs), methylation, and fragmentomics (*3*). Large clinical trials have demonstrated the promise of liquid biopsy as noninvasive Multi-Cancer Early Detection tests (MCEDs), but performance remains limited for early-stage cancers and even poorer for precancerous lesions (*4–8*). These limitations are primarily driven by the extremely low abundance of ctDNA at low tumor burdens and the overwhelming background of normal cell-free DNA (cfDNA). Importantly, not all precancers necessitate clinical interventions (*9*). An ideal screening test would therefore not only detect lesions at the pre-malignant stage but also enable risk stratification, identifying those at elevated risk for progressing to cancer while sparing low-risk individuals from unnecessary medical procedures and anxiety. However, most existing liquid biopsies are limited to the binary detection of ctDNA signals, rather than providing informative risk stratification necessary in screening settings (*8*, *10*).

Human papillomavirus (HPV) causes six cancer types, which together account for ∼5% of all cancers worldwide, with HPV-associated oropharyngeal, cervical, and anal cancers being the most prevalent (*11–13*). Progression from HPV infection to cancer spans decades, offering a valuable window for early detection (*11*). Targeting etiologic HPV cfDNA in blood provides a unique opportunity to overcome the key technical constraints in liquid biopsy. Since HPV is not a bloodborne pathogen, HPV cfDNA in blood should therefore originate from pathologic cell turnover and is inherently specific to the lesion sites. In addition, the lack of sequence homology between HPV and human genomes enables sensitive HPV cfDNA detection against the human background. Furthermore, HPV-infected cells can harbor tens to thousands of copies of the viral genome, providing an intrinsic sensitivity boost (*14*, *15*). We have previously developed a multi-feature liquid biopsy, HPV-DeepSeek, which uses hybrid-capture enrichment against the HPV whole genome across 42 genotypes and selected human genomic regions for targeted deep sequencing (*16*, *17*). The assay previously achieved detection of HPV cfDNA in biobanked plasma samples collected years before clinical diagnosis of HPV+ oropharyngeal cancer, establishing the feasibility of blood-based HPV+ cancer screening (*17*).

The presence of HPV cfDNA years before cancer diagnosis raises the possibility that in addition to cancer, HPV precancer could be detected from blood. Here, we use HPV+ anal cancer as the model system to examine blood-based detection and risk stratification of HPV precancer and cancer. We chose HPV+ anal cancer due to its well-defined precancer stages and the urgent clinical need for effective screening. Anal cancer, although once considered rare, has seen a steady rise in incidence over the past three decades, particularly among high-risk populations such as people living with HIV and men who have sex with men (*18–20*). Nearly all anal cancers are attributable to HPV (*19*). The recent Anal Cancer-HSIL Outcomes Research (ANCHOR) trial demonstrated the clinical benefits of treating anal high-grade precancers, underscoring the need for screening (*21*), and the HIV Medicine Association and the International Anal Neoplasia Society now recommend anal cancer screening in high-risk populations (*22*). However, current tissue-based screening approaches, namely cytology and HPV testing, are invasive, require specialized infrastructure, and lack clinical specificity (*23*). Furthermore, the diagnostic workup following abnormal screening, high-resolution anoscopy (HRA), is in severe shortage, highlighting the urgent need for novel strategies with high accuracy for anal cancer and precancer screening (*24–26*).

In this prospective observational study, we applied HPV-DeepSeek to plasma samples spanning the full anal HPV infection-precancer-cancer continuum, to systematically evaluate blood-based detection of HPV cfDNA and genomic features linked to malignant risk. Leveraging the tumor-specific origin of HPV cfDNA, we further performed fragmentomics analysis to identify additional stage-specific signatures. Ultimately, we developed a unified classifier incorporating all assay features to detect cancer and high-grade precancer, stages in need of clinical intervention.

## RESULTS

### Study design and HPV-DeepSeek liquid biopsy framework

In anal cancer, HPV infection progresses through anal intraepithelial neoplasia 1 (AIN1), AIN2, and AIN3, to eventual invasive anal squamous cell carcinoma (ASCC) (**Figure 1A**). We assembled a cohort of plasma samples covering the entire continuum, comprising biobanked samples collected prior to ASCC diagnosis (n=6) and prospectively collected samples during routine anal cancer screening (n=104) and from general population controls (n=60). We applied HPV-DeepSeek, a custom liquid biopsy employing targeted HPV whole-genome sequencing for ultrasensitive HPV cfDNA detection (**Figure 1B**). HPV-DeepSeek enabled comprehensive profiling of stage-informative molecular features, including HPV genotype, HPV structural variations (HPV-HPV rearrangement and HPV-human integrations), human CNAs, *PIK3CA* mutations, and fragmentomics signatures, which were integrated into a unified classifier to detect ASCC, AIN3, and AIN2, stages that require clinical treatment (**Figure 1C**).

**Figure 1.**
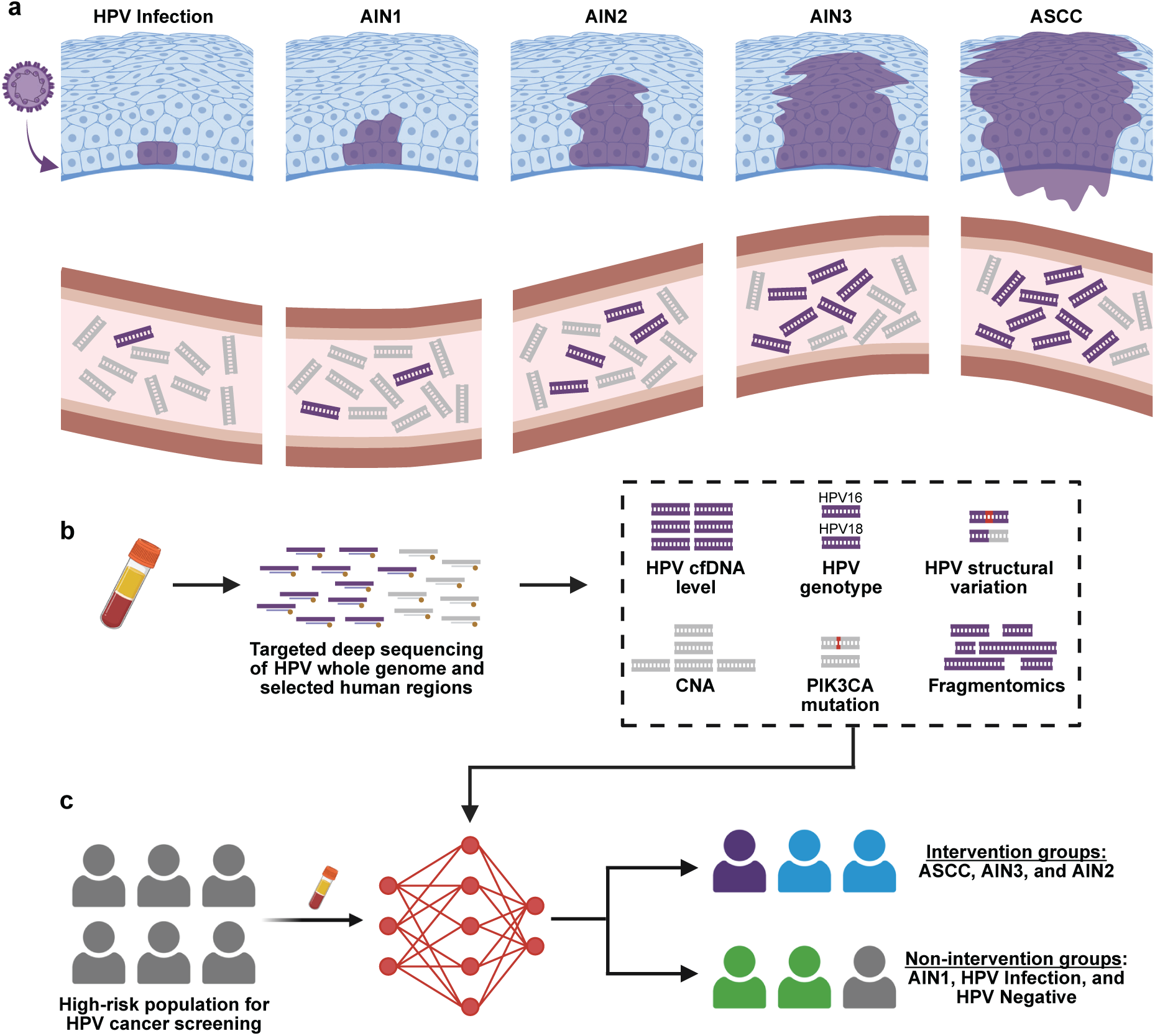
Study design for blood-based detection and molecular risk stratification of HPV-associated cancer and precancer. (**A),** In anal cancer, HPV infection progresses through well-defined precancer stages, including anal intraepithelial neoplasia 1 (AIN1), AIN2, and AIN3, to invasive anal squamous cell carcinoma (ASCC). Blood HPV cell-free (cf)DNA originating from cell turnover is lesion-specific and readily distinguishable from the human cfDNA background, offering a unique opportunity to overcome current liquid biopsy constraints. (**B),** Schematic of HPV-DeepSeek, a custom multi-feature liquid biopsy for HPV cfDNA. Plasma cfDNA undergoes targeted deep sequencing using hybrid-capture probes spanning HPV whole genomes across 42 genotypes and selected human genomic regions. Multiple features are derived for detection and risk stratification, including HPV cfDNA level, HPV genotype, HPV structural variations (HPV-HPV rearrangement and HPV-human integration events), human copy number alterations (CNAs), *PIK3CA* mutations, and HPV cfDNA fragmentomics signatures. (**C),** A unified classifier incorporating all assay features addresses a critical unmet need in anal cancer screening by identifying ASCC, AIN3, and AIN2, stages requiring clinical intervention.

### HPV cfDNA is detectable years before the clinical diagnosis of anal cancer

We first investigated whether HPV cfDNA could be detected in plasma prior to clinical diagnosis of anal cancer. In the Mass General Brigham Biobank, six cases were identified who were diagnosed with ASCC, each with one pre-diagnostic plasma sample available collected more than one year prior to ASCC diagnosis (range: 2.8–8.6 years; median lead time: 4.5 years) (**Supplementary Table 1**). Using the previously established assay threshold, four of six (66.7%) pre-diagnostic plasma samples tested positive for HPV cfDNA, with a maximum detection lead time of 7.2 years (**Figure 2A and Supplementary Figure 1**) (*16*, *17*). Three positive cases were genotype HPV16 and one was HPV33. Tissue biopsy samples obtained at the time of cancer clinical diagnosis were available for two cases and demonstrated concordant HPV genotypes with the corresponding pre-diagnostic plasma samples (both HPV16).

**Figure 2.**
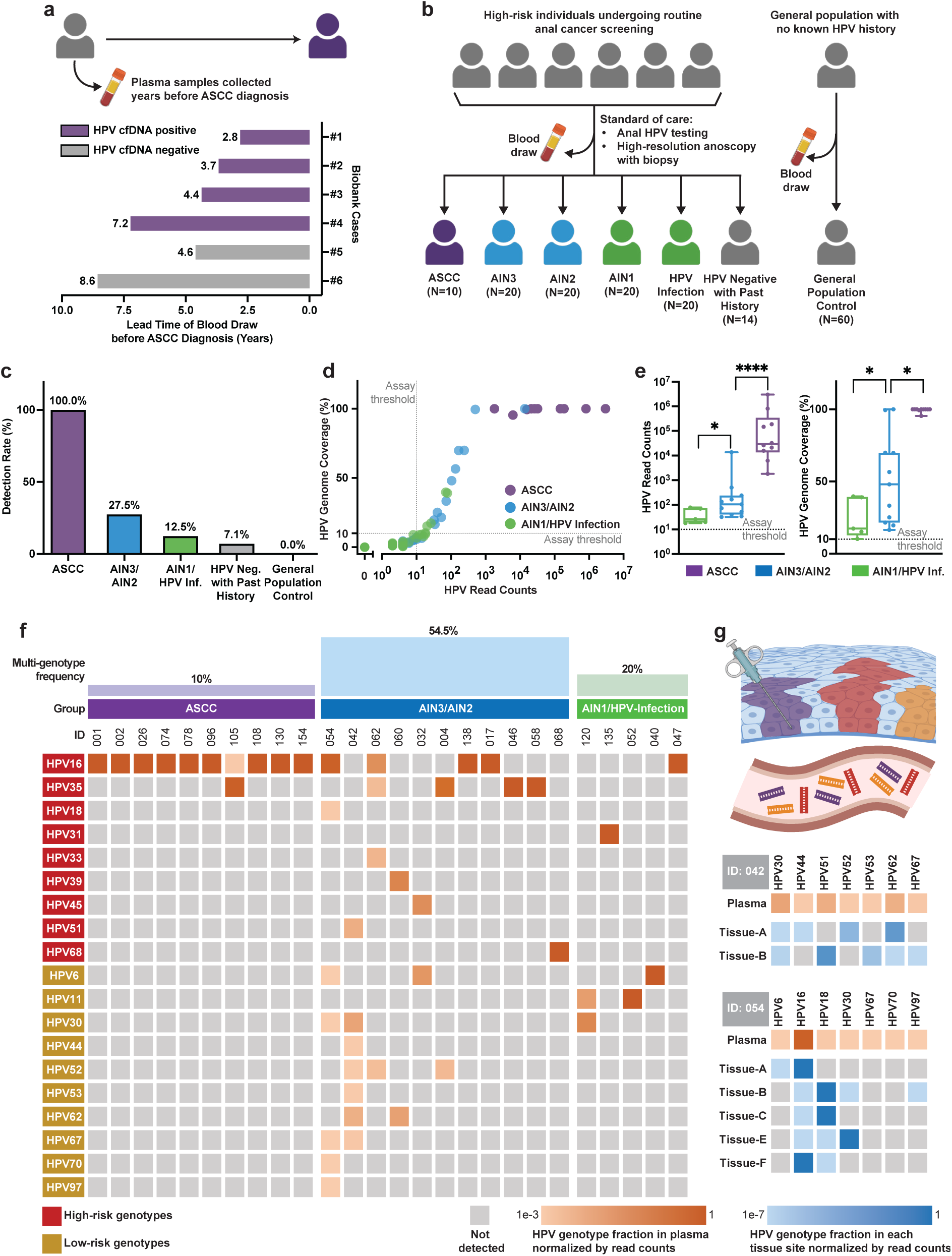
Detection of HPV cfDNA along the infection-precancer-cancer continuum. **(A),** Detectability of HPV cfDNA in biobanked plasma samples collected years before clinical diagnosis of ASCC. **(B),** Overview of the sample collection in the prospective screening cohort. (**C),** Detection rates of HPV cfDNA across the clinical groups in the prospective cohort. (**D),** Primary assay outputs, HPV cfDNA levels, represented by HPV read counts and fraction of the HPV genome detected (referred as HPV genome coverage), for samples across the HPV infection-precancer-cancer continuum. The assay threshold was established previously through analytical characterization. (**E),** Comparison of HPV cfDNA levels among positive samples across disease stages. Boxes indicate the interquartile range with the median shown as the central line, and whiskers extend to the minimum and maximum values. Statistical significances were determined using a one-sided Wilcoxon rank-sum test (\**p* < 0.05; \*\*\*\**p* < 0.0001). (**F),** HPV genotype distributions among cases positive for HPV cfDNA, with the bar plot showing the frequency of multi-genotype cases within each disease stage. (**G),** Concordance between HPV genotypes detected in plasma and matched tissue biopsies in two multi-genotype cases. For each case, multiple tissue biopsies were sampled during standard clinical care. For patient ID 054, the sample Tissue-D was not available for this study.

### HPV cfDNA is detectable beginning at the infection stage, with increasing levels as the stages progressed

We next prospectively collected blood samples from high-risk population undergoing routine anal cancer screening (**Figure 2B and Supplementary Table 2**). This screening cohort comprised individuals at elevated risk for anal cancer, including people living with HIV (PLWH) and HIV-negative individuals with a prior history of treated HPV precancerous lesions. We chose this cohort to mimic the real-world screening utility of the developed liquid biopsy approach. Patients were classified based on clinical tissue biopsy diagnosis, including anal HPV infection, AIN1, AIN2, AIN3, and ASCC. Clinically, it is critical to distinguish high-grade lesions (AIN3 and AIN2) requiring intervention from low-grade lesions (AIN1) or HPV infection that can be safely monitored. Accordingly, AIN3 and AIN2 were grouped for analysis, as were AIN1 and HPV infection. Patients in the high-risk cohort but negative for clinical HPV testing were classified as anal HPV-negative. Additionally, we collected blood samples from the general population controls with no known HPV history.

General population controls with no known HPV history were all negative for HPV cfDNA, demonstrating 100% analytical specificity (0/60; 95% CI: 95.1–100%, exact binomial) of the HPV-DeepSeek assay (**Figure 2C**). All ASCC cases were positive for HPV cfDNA, demonstrating 100% sensitivity (10/10; 95% CI: 74.1–100%, exact binomial) for blood-based anal cancer detection. In the precancerous stages, HPV cfDNA was detectable beginning at the HPV infection stage, with higher detection rates in AIN3/AIN2 (27.5%) compared with AIN1/HPV infection (12.5%). Among the positive samples, the HPV cfDNA level, quantified by HPV read counts and HPV genome coverage, increased significantly with the disease severity (**Figure 2D and E**). Median HPV read counts in ASCC, AIN3/AIN2, and AIN1/HPV infection were 29163, 104, and 26, respectively, and the median HPV genome coverage was 100%, 48%, and 17%. One sample in the anal HPV-negative group was positive for HPV cfDNA. Interestingly, the patient was later clinically diagnosed with anal HPV infection at one-year follow-up.

### HPV genotypes show distinct distributions across cancer and precancer stages

HPV genotypes among positive samples exhibited distinct distributions in cancer versus precancer stages (**Figure 2F**). In ASCC, HPV16 was the dominant genotype and detected in all cases, with one sample harboring additional HPV35. In contrast, precancer stages showed diverse genotypes, with a total of 19 genotypes detected, including 10 low-risk genotypes. Consistent with previous tissue-based studies of cervical and anal precancers, multi-genotype infections were frequently observed in the precancer stages, occurring in 54.5% of AIN3/AIN2 positive samples and 20% of AIN1/HPV infection (**Figure 2F and Supplementary Figure 2**) (*27–29*). Samples harbored up to seven distinct genotypes. The most frequent genotypes detected in precancer stages were HPV16 (31.3%), HPV35 (25.0%), HPV6 (18.8%), HPV30 (18.8%), and HPV52 (18.8%). Notably, while all samples in AIN3/AIN2 contained at least one high-risk genotype, three of the five (60%) samples in AIN1/HPV infection detected only low-risk genotypes, demonstrating a significantly higher frequency of low-risk-only infections in AIN1/HPV infection compared with AIN3/AIN2 (**Supplementary Figure 3**).

### HPV cfDNA uniquely captures inter-lesional heterogeneity

For the two samples with the highest number of HPV genotypes detected in blood, we further sequenced the paired tissue biopsy samples using HPV-DeepSeek to compare with plasma findings (**Figure 2G**). Multiple lesion sites were sampled for each patient as part of standard clinical care. In the first case (ID: 042), each lesion site harbored a subset of the HPV genotypes found in blood, which collectively recapitulated the full genotype spectrum found in HPV cfDNA. In the other case (ID: 054), similarly, each lesion site contained a subset of the blood HPV genotypes. Two genotypes detected in blood (HPV67 and HPV70) were not present in the available tissue samples; however, one lesion site was not available for sequencing. The complementary distribution of HPV genotypes across multiple tissue biopsy sites underscores the need for multi-site tissue sampling during anal cancer screening, consistent with prior observations in cervical cancer screening (*30*). Conversely, the detection of HPV genotypes originating from multiple lesion sites within a single blood sample highlights the potential of blood-based analysis to overcome tissue sampling bias inherent to multi-site disease processes.

### HPV integration and rearrangement events differentiate cancer and non-cancer cases

In infected cells, HPV genomes typically exist as episomes; however, integration of HPV DNA into the human genome, as well as rearrangements within HPV genomes, are recognized genomic hallmarks in HPV carcinogenesis (*31–33*). We previously demonstrated the ability of HPV-DeepSeek to accurately detect and classify HPV integration and rearrangement events from blood in HPV-associated oropharynx cancer (*34*). Here we applied the same strategy to the screening cohort aiming to improve cancer risk stratification (**Figure 3A**). HPV integration or rearrangement events were identified in 9 out of the 10 cancer cases (90%). 3 cases had HPV-HPV rearrangements only, 5 had mixed HPV-human integration and HPV-HPV rearrangements, and 1 had clonal HPV-human integration (**Figure 3B**). In contrast, no HPV integration or rearrangements were detected in the precancer or control samples.

**Figure 3.**
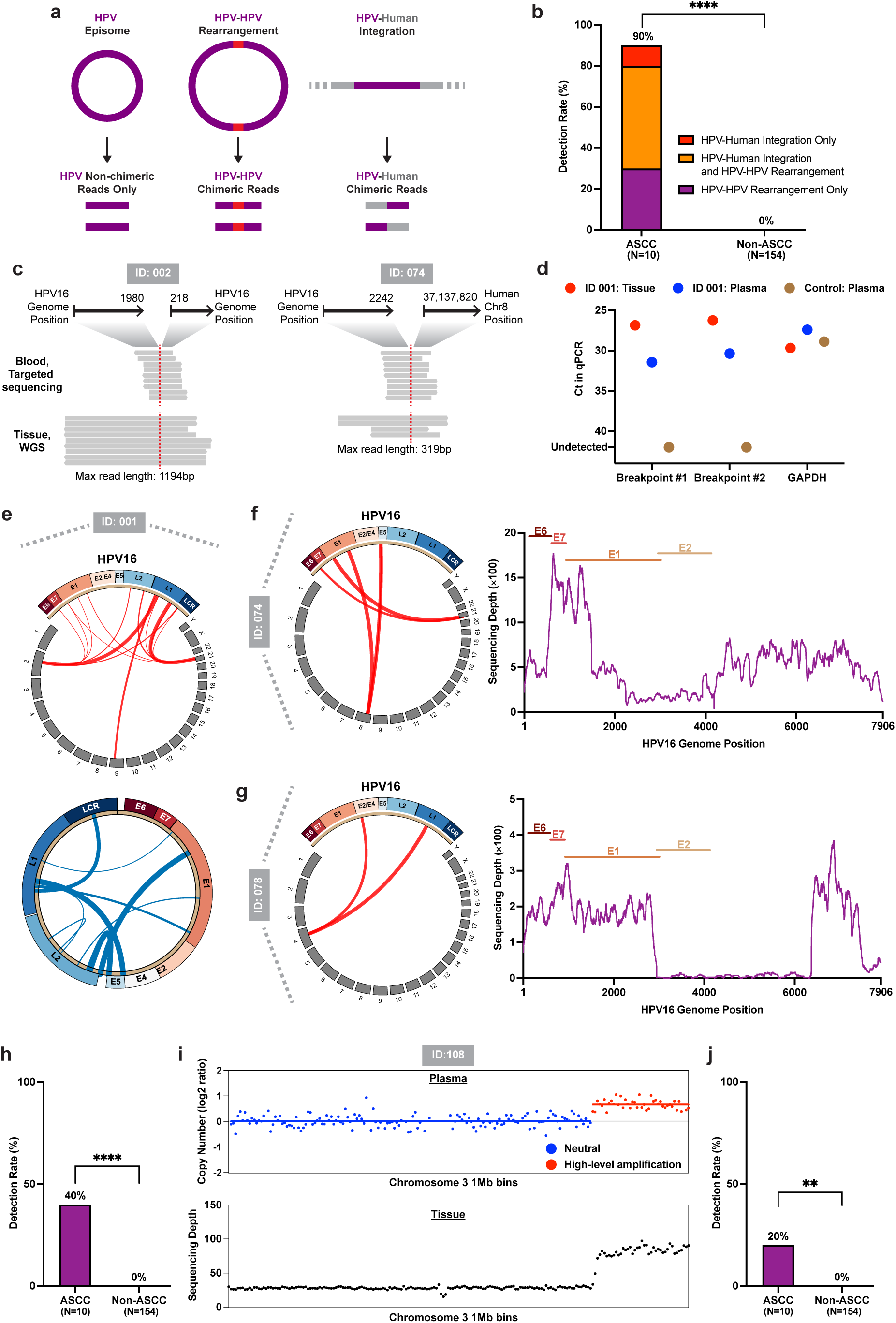
Blood-based detection of additional genomic hallmarks in HPV malignancy. (**A),** Schematic illustrating using HPV-HPV and HPV-human chimeric reads to detect HPV-HPV rearrangements and HPV-human integration events, respectively. (**B),** Detection rates of HPV structural variations in the cancer versus non-cancer cases. (**C),** Examples of long-read WGS of paired FFPE tissue samples validating two events identified in plasma. (**D),** PCR validation of two breakpoints found in one sample. Note that lower Ct in qPCR represents higher counts of target DNA (y axis). (**E),** Circos plots showing representative case with concurrent complex HPV–human integration events (top) and HPV–HPV rearrangements (bottom) detected by HPV-DeepSeek. The HPV genome is shown as a sector annotated with viral genes, with human chromosomes shown as gray sectors. Red links indicated HPV–human breakpoints and blue links indicate HPV-HPV non-circular junctions. Link width scaled with the number of supporting reads. (**F),** Representative case illustrating HPV16–human integration associated with loss of *E1*/*E2* and relative amplification of *E6*/*E7*. (**G),** Representative case showing clonal HPV16 integration with *E2* deletion. (**H),** Detection rates of 3q amplification in cancer versus non-cancer cases. **(I)** Representative case demonstrating concordance between blood-based detection of 3q amplification and WGS depth of the paired tissue sample. (**J),** Detection rates of *PIK3CA* mutations in cancer versus non-cancer cases. Statistical significance in panel b, h, and j was determined using Fisher’s exact test (\*\**p* < 0.01, \*\*\*\**p* < 0.0001).

To further confirm the accuracy of HPV integration and rearrangement calling from our assay in this cohort, we conducted long-read whole-genome sequencing (WGS) on 4 paired tissue samples from ASCC cases with sufficient DNA. Tissue WGS validated one breakpoint identified in the matched plasma samples in each case (**Figure 3C and Supplementary Figure 4**). Additional breakpoints found in plasma liquid biopsy but not in tissue WGS could be due to limited sequencing depth of the HPV genome in WGS or the compromised FFPE DNA quality. To further assess breakpoints not confirmed by tissue WGS, we performed PCR validation of two breakpoints found in one plasma sample (ID 001). PCR assays designed to specifically amplify chimeric reads spanning the breakpoints showed successful detection in the matched plasma and tissue DNA samples, for both breakpoints, while control plasma samples lacking the targeted chimeric reads showed no PCR amplification, using comparable DNA input as verified by a control PCR assay targeting the human *GAPDH* gene (**Figure 3D**).

Notably, the comprehensive characterization of chimeric reads in our assay revealed a diverse and complex landscape of HPV integration and rearrangement in ASCC, consistent with known mechanisms of HPV carcinogenesis (**Supplementary Figure 5**). For example, one case exhibited multiple concurrent events of both HPV-human integration and HPV-HPV rearrangements (**Figure 3E**). While HPV breakpoints occurred across the viral genome, the human breakpoints clustered within a few regions, consistent with prior reports of HPV integration inducing focal genomic instability with recurrent integration events (*33*, *35*). In addition, HPV integration led to the deletion of the viral *E1* and *E2* genes (**Figure 3F**), including one case demonstrating clonal integration (**Figure 3G**), consistent with previous findings of integration disrupting HPV *E1* and *E2*, known regulators of oncoprotein E6 and E7 expression (*31*).

Considering the frequent detection of HPV integration or rearrangement among ASCC cases and their association with escalated cancer risk, we next investigated the biobanked pre-diagnostic cohort (**Figure 2A**). An HPV-human integration event was identified in one plasma sample collected 4.4 years prior to clinical cancer diagnosis. Sequencing of the FFPE tissue block at the time of cancer diagnosis validated the same integration event in the tumor (**Supplementary Figure 6**). Together, the exclusive detection of HPV integration or rearrangement in cancer cases within the prospective cohort, and its presence in pre-diagnostic plasma from a patient who was later diagnosed with cancer, highlighted the potential of HPV integration and rearrangements as biomarkers for improved cancer risk stratification.

### Cancer-associated *PIK3CA* mutations and 3q amplification are detectable in blood exclusively in cancer cases

In addition to characteristic features in the HPV genome, we next assessed the detection of human genomic hallmarks associated with HPV+ cancers across the developmental stages. We focused on 3q amplification and *PIK3CA* mutations as they were among the most common genomic alterations in HPV+ cancers (*36*, *37*).

Recurrent amplification of 3q, the region harboring key oncogenic genes in squamous cell carcinoma including *PIK3CA, TP63,* and *SOX2*, is the most frequent and characteristic CNA event in HPV+ cancers. Leveraging the off-target reads from HPV-DeepSeek distributed throughout the human genome as ultra-low-pass WGS data, we applied ichorCNA to detect 3q amplification from plasma cfDNA (*38*). Because the off-target reads yielded WGS coverage below the recommended range for tumor fraction estimation by ichorCNA (median, 0.05x; range, 0.01-0.23x), we restricted the analysis to detection of the established 3q amplification to improve specificity. 4 out of the 10 cancer cases (40%) had 3q amplification detected (**Figure 3H**), aligning with the reported 50-60% rates in tissue-based studies (*39*, *40*). Paired tissue biopsy WGS data were available for two cases which validated the presence of 3q gains (**Figure 3I and Supplementary Figure 7**). No 3q amplification were detected in the non-ASCC samples.

*PIK3CA* mutations were detected in two ASCC cases (20%), consistent with reported 22% frequency in tissue-based studies (**Figure 3J**) (*41*). One case had paired tissue WGS data available finding the concordant mutation. No *PIK3CA* mutations were detected in the non-ASCC samples.

### HPV cfDNA fragmentomics exhibits progressive changes along the infection-precancer-cancer continuum

Given prior evidence that ctDNA fragments differ in length from healthy cfDNA (*42*) and that viral cfDNA can exhibit distinct fragmentation characteristics reflecting viral biology and tumor state (*43–45*), we next performed fragmentomics analysis of HPV cfDNA, leveraging its lesion-specific origin to identify additional features that could further improve risk stratification. Because HPV cfDNA detection rates were low in the AIN1/HPV-infection group, an additional four patients clinically diagnosed with AIN1/HPV infection without tissue biopsy were included for fragmentomics signature discovery.

In ASCC, HPV cfDNA displayed distinct length profiles compared with human cfDNA from the same samples, with overall shorter fragments characterized by a mono-nucleosomal peak at 143 bp versus 167 bp in human cfDNA, as well as a left-shifted and attenuated di-nucleosomal peak (**Figure 4A**), consistent with prior reports (*43*). The distinct length profile of HPV cfDNA may reflect differences in nucleosome organization between the HPV and human genomes, as well as the tumor-derived origin of HPV cfDNA while human cfDNA predominantly originates from healthy cells.

**Figure 4.**
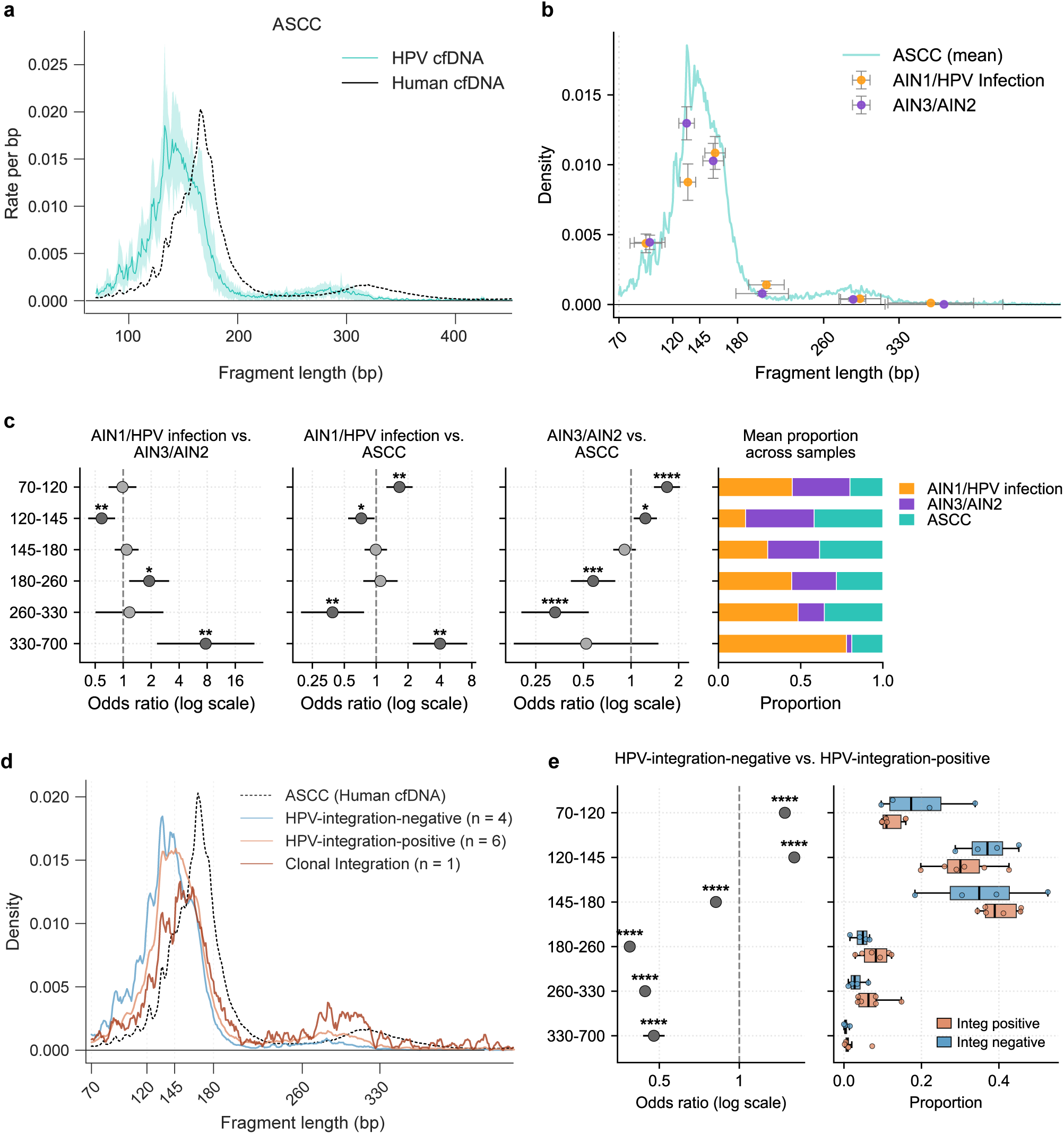
HPV cfDNA fragmentomics signatures. **(A),** Fragment length distributions of cfDNA across disease states. Fragment length profiles are shown as rate per base pair for HPV cfDNA (colored) and human cfDNA (black) in ASCC samples (n=10). Solid lines represent the mean fragment-length distribution across samples, and shaded bands indicate the 10th–90th percentile range of the HPV cfDNA distribution. (**B),** Differential fragment length enrichment of HPV cfDNA in precancerous lesions and anal cancer. Fragment length distributions of HPV cfDNA are shown as density per base pair for AIN1/HPV infection (n=24) and AIN3/AIN2 (n=20) overlaid on the mean anal ASCC fragment-length profile. Points represent the average rate per base pair within each coarse fragment-length bin, plotted at the bin center of mass. Horizontal error bars reflect fragment-length dispersion (standard deviation) within each bin, while vertical error bars indicate the standard error of the estimated rate per base pair. **(C),** Fragment-length bin enrichment across disease stages. Forest plots showing odds ratios (ORs) and 95% confidence intervals for enrichment of HPV-derived cfDNA fragments within each coarse fragment-length bin (70–120, 120–145, 145–180, 180–260, 260–330, 330–700 bp). ORs were calculated using Fisher’s exact test on aggregated fragment counts (bin vs. non-bin) for three pairwise comparisons: AIN1/HPV infection vs. AIN3/AIN2 (left), AIN1/HPV infection vs. ASCC (middle), and AIN3/AIN2 vs. ASCC (right). Odds ratios are plotted on a log scale with the dashed vertical line indicating OR = 1. Asterisks denote significance after Benjamini–Hochberg multiple-testing correction across bins (\**q* < 0.05, \*\**q* < 0.01, \*\*\**q* < 0.001, \*\*\**q* < 0.0001). Stacked bar plots on the far right show the mean proportion of fragments per length bin attributed to each stage, normalized across groups. (**D),** Integration-specific length profiles of HPV cfDNA in ASCC samples. Fragment length distributions of HPV cfDNA in ASCC samples stratified by viral integration status: integration-negative (n = 4) and integration-positive (n = 6), with one sample exhibiting clonal integration. The mean human cfDNA fragment-length profile from ASCC (same as in panel A) is shown with a dashed line as a reference. **(E),** Integration-associated fragment-length enrichment in ASCC. Forest plot *(Left)* showing fragment-length bin enrichment comparing integration-positive versus integration-negative ASCC samples. Odds ratios and 95% confidence intervals were calculated from Fisher’s exact test on aggregated fragment counts (bin vs. non-bin) and are displayed on a log scale (dashed line, OR = 1). Asterisks indicate significance after Benjamini–Hochberg correction across bins (\*\*\*\**q* < 1e−4). Horizontal boxplots *(Right)* showing the per-sample proportion of fragments within each fragment length bin for HPV integration-positive (orange) and integration-negative (blue) samples. Boxes span the interquartile range (IQR), with the vertical line indicating the median; whiskers extend to 1.5 × IQR. Individual sample values are overlaid as dots.

Comparing HPV cfDNA length profiles among AIN1/HPV infection, AIN3/AIN2, and ASCC groups revealed progressive changes along the HPV infection-precancer-cancer continuum (**Figure 4B** and **Supplementary Figure 8A**). Due to the small number of HPV cfDNA fragments in precancer samples, fragment lengths were summarized into six bins (70-120, 120-145, 145-180, 180-260, 260-330, and 330-700 bp), and the density of fragments were compared to ASCC distribution. Notably, the AIN1/HPV infection group showed depletion of mono-nucleosomal peaks (120-145 bp) compared with more advanced stages (**Figure 4B**), consistent with less defined nucleosomal organization in early-stage disease. AIN3/AIN2 manifested an intermediate profile, with a mono-nucleosomal peak approaching ASCC but still lacking di-nucleosome peak formation.

To quantify these shifts, we performed pairwise comparisons using aggregated HPV cfDNA fragment counts per group. Two AIN3/AIN2 outliers with unusually high HPV fragment counts were excluded to prevent disproportionate influence on the aggregated spectrum (**Supplementary Figure 8B**). When comparing the two precancer groups, AIN1/HPV infection samples were significantly depleted (OR=0.59) of mono-nucleosomal fragments (120-145 bp) and enriched in longer fragments, particularly in the 330-700 bp range (OR=7.69) (**Figure 4C**). Compared with ASCC, both precancer groups exhibited depletion (OR=0.39 for AIN1/HPV infection; OR=0.33 for AIN3/AIN2) in the di-nucleosomal peak (260–330 bp) and enrichment of the shortest fragments (70–120 bp) (**Figure 4C**). These differences may reflect underlying changes in chromatin structure and viral genome organization, suggesting increasingly defined nucleosomal fragmentation patterns with advancing disease stages.

### HPV integration drives the emergence of the di-nucleosome peak in HPV cfDNA

Given the observed enrichment of di-nucleosomal fragments in ASCC, we hypothesized that HPV integration into host chromatin may contribute to this shift in fragmentation pattern. We therefore examined HPV cfDNA fragmentation profiles stratified by HPV integration status within ASCC. Integration-positive cases exhibited fragment length distributions more similar to human cfDNA, characterized by a longer mono-nucleosomal peak and a more pronounced di-nucleosomal peak, and this pattern was particularly evident in the ASCC case with clonal integration (**Figure 4D**). Analysis across fragment-length bins further confirmed significant shifts in fragment proportions between integration-positive and integration-negative samples (**Figure 4E**). These findings are consistent with the notion that integrated HPV genomes, embedded within host chromatin, may undergo nucleosome-associated fragmentation similar to human DNA, thereby generating viral cfDNA fragment profiles that closely resemble human cfDNA patterns.

### Unsupervised clustering reveals distinct HPV cfDNA fragmentomic subtypes across disease stages

Although the aggregated fragment length profiles showed gradual shifts across stages (**Figure 4C**), they do not capture potential inter-sample variability. Hypothesizing that each disease stage could harbor heterogenous fragmentomics subtypes, we performed unsupervised clustering of HPV cfDNA fragment length profiles within each disease stage. To compare the fragmentomics subtypes across disease stages, we then conducted a second round of clustering using the average profiles of the initial clusters (**Supplementary Figure 9)**. We identified three major fragmentomics patterns encompassing the majority of samples. The first was dominated by short fragments (70–120 bp) and consisted exclusively of precancer samples, with enrichment from AIN1/HPV-infection. The second exhibited a prominent peak at 145–180 bp, consistent with mono-nucleosomal fragments, and included both precancer and cancer samples with enrichment in ASCC cases. The third showed a peak at 120–145 bp and comprised AIN3/AIN2 and ASCC samples, with no representation from AIN1/HPV-infection. In addition to these three predominant patterns, two minor clusters specific to the AIN1/HPV-infection stage exhibited longer fragment distributions that did not correspond to canonical mono- or di-nucleosomal peaks.

Together, these findings demonstrated that HPV cfDNA fragmentomics harbored stage-informative features that could further improve blood-based risk stratification. While the clusters showed enrichment by disease stages, they were not strictly stage-specific, with overlapping fragmentation patterns observed across groups. This suggests that HPV cfDNA fragmentation patterns evolve progressively, with transitional profiles shared between adjacent disease stages.

### An integrated classifier differentiates lesions which require clinical intervention from those that can be observed

Leveraging our ability to detect HPV cfDNA across the infection-precancer-cancer continuum and the molecular and fragmentomics features stratifying developmental stages, we aimed to build an integrated classifier to identify anal cancer and high-grade precancerous lesions (AIN3/AIN2), stages that require clinical intervention. To evaluate the potential clinical utility in a real-world screening setting, we restricted model training and validation to the high-risk screening cohort and excluded the general population controls, as this cohort represented individuals likely to be screened for anal cancer and precancer based on current guidelines.

For fragmentomics features, to accommodate the identified inter-sample variability and the sparse HPV cfDNA fragment counts in precancer stages, we employed a likelihood-based approach to annotate sample-level fragmentomics length profiles. Specifically, for each sample, we calculated the likelihood of assignment to each of the 9 fragmentomics clusters (**Supplementary Figure 9**). We then summed the likelihoods of clusters corresponding to each disease stage, generating stage-specific likelihoods that were used as the input features in the classifier (**Figure 5A**).

**Figure 5.**
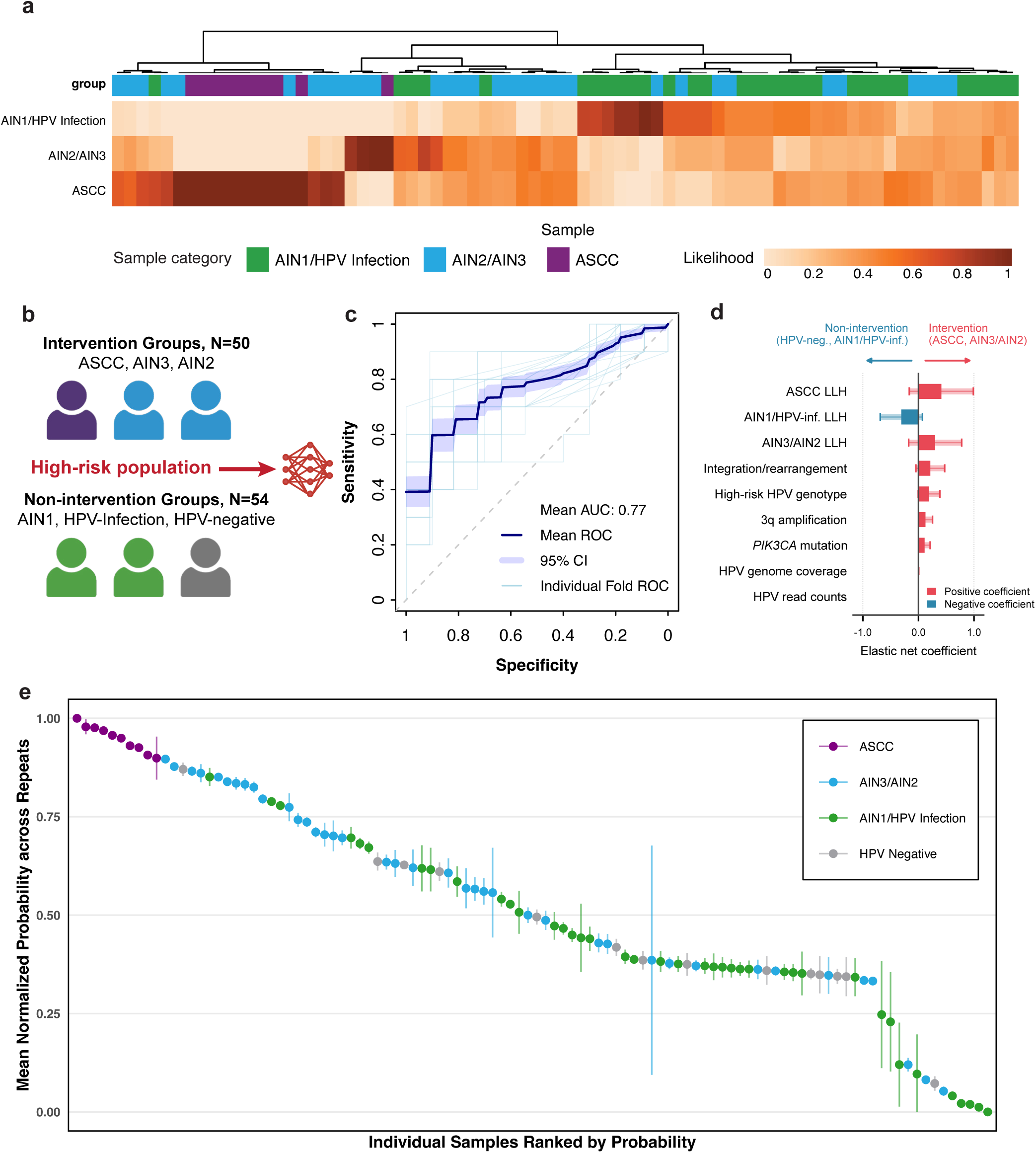
Integrated classifier for detecting HPV cancer and high-grade precancer in need of clinical interventions. **(A),** Clustering of samples in the high-risk screening cohort based on fragmentomics-derived stage-specific likelihoods. Each column represents an individual sample within the prospective high-risk screening cohort, and each row corresponds to their fragmentomics likelihoods to AIN1/HPV-infection, AIN2/AIN3, or ASCC profiles. (**B),** Overview of the sample cohort used to develop the classifier. (**C),** Classifier performance for detecting intervention groups (ASCC, AIN3, and AIN2) in the high-risk screening cohort, evaluated using repeated nested 5-fold cross-validation. The mean ROC across all folds, the 95% confidence interval of the mean ROC, and the ROC of each individual fold are shown. (**D),** Feature coefficients of the integrated classifier across cross-validation splits. ASCC LLH, AIN3/AIN2 LLH, and AIN1/HPV-inf. LLH represent fragmentomics-derived likelihoods to ASCC, AIN3/AIN2, and AIN1/HPV-infection stages, respectively. Horizontal bars show the mean elastic net coefficient across 25 nested cross-validation folds for each feature, with error bars and shaded bands indicating ± standard deviation. Positive coefficients indicate association with intervention groups while negative coefficients indicate association with non-intervention groups. Features are ordered by absolute mean coefficient magnitude. **(E),** Predicted probabilities of assignment to the intervention group by the classifier for each sample. Dots represent the mean normalized probabilities across the five repeats of the classifier cross-validation, and vertical lines indicate the 95% confidence interval.

Incorporating all the informative features, including HPV cfDNA level, presence of high-risk HPV genotype, HPV-human integration or HPV-HPV rearrangements, 3q amplification, *PIK3CA* mutations, and the fragmentomics stage-specific likelihoods, we developed a logistic regression model for classifying samples into intervention groups (ASCC, AIN3, and AIN2) and non-intervention groups (AIN1, HPV-infection, and HPV-negative) (**Figure 5B**). Using repeated nested cross validation, the model achieved a mean AUC of 0.77 (95% CI: 0.742-0.797), with a mean sensitivity of 73.2% (95% CI: 68.9%-77.5%) and a mean specificity of 82.7% (95% CI: 78.6%-86.7%) at the Youden index threshold (**Figure 5C**). Feature analysis across folds showed consistent selection of all the features in most folds with stable coefficient directions, supporting the robustness of the classifier despite the modest sample size (**Figure 5D**). The stage-specific fragmentomics likelihoods were the most informative features across the splits.

While HPV high-grade precancers require clinical treatment, not all are progressive, with studies in cervical cancer estimating only ∼30% progress to invasive cancer if untreated, indicating biological heterogeneity within HPV high-grade precancers (*46*, *47*). To explore whether the classifier could capture underlying biological cancer risk, we evaluated sample-level predictions across validation folds. ASCC cases showed the highest predicted probabilities and were consistently classified as positive (intervention group) across all folds using the Youden index threshold, demonstrating the capability of the classifier in detecting cancer-like signals (**Figure 5E and Supplementary Figure 10A**). High-grade precancer samples exhibited heterogenous prediction patterns, with some consistently assigned high probabilities and some consistently low. 40% of high-grade precancers were classified as positive across all folds, and 32.5% were consistently classified negative, supporting the heterogeneity within high-grade precancers. The finding was particularly pronounced when only considering the features of HPV cfDNA read counts and the fragmentomics AIN1/HPV-infection likelihood (**Supplementary Figure 10B**). While ASCC and AIN1/HPV-infection samples occupied the opposite distinct ends of this feature space, AIN3/AIN2 samples were distributed between the extremes, with some clustering closer to AIN1/HPV-infection and others aligning more closely with cancer-like profiles. These observations raise the question of whether the subset of AIN3/AIN2 high-grade precancers with scores closest to the cancer group may have a different risk of progression compared to those with lower scores. However, validation would require longitudinal follow-up without immediate clinical intervention.

## DISCUSSION

Liquid biopsies offer an attractive strategy for cancer screening, yet current performance remains limited for detecting early-stage cancer and precancer, due to the extremely low abundance of ctDNA amid an overwhelming background of cfDNA. In HPV-associated cancers, HPV cfDNA offers a unique opportunity to overcome current liquid biopsy constraints, owing to its lesion-specific origin, lack of sequence homology to human DNA, and the intrinsic sensitivity boost conferred by multiple viral genome copies per cell. In this study, we applied a custom, multi-feature liquid biopsy targeting the HPV whole genome and selected human regions and demonstrated the potential for blood-based detection and molecular risk stratification across the HPV infection-precancer-cancer continuum.

We first applied the custom liquid biopsy to biobanked plasma samples collected prior to ASCC clinical diagnosis, finding that HPV cfDNA was detectable more than seven years before the clinical diagnosis. These results demonstrate that blood-based screening for anal cancer is feasible. Together with our previous study in HPV-associated oropharyngeal cancer, the findings highlight the broad potential of HPV cfDNA for pan-HPV cancer screening (*17*).

Next in the prospective screening cohort spanning the full HPV infection-precancer-cancer continuum, we demonstrated that HPV cfDNA was detectable as early as the infection stage, the inception of HPV cancer development, with increasing detection rates and HPV cfDNA levels as stages progressed. Compared to prior studies using droplet digital PCR (ddPCR) reporting limited to no detection of blood HPV cfDNA during precancer stages, HPV-DeepSeek employs a hybrid-capture probe panel spanning the entire HPV genomes across 42 common genotypes for targeted deep sequencing, enabling detection of fragmented HPV cfDNA throughout the viral genome (*48*). Indeed, we observed a median HPV genome coverage of 48.0% in AIN3/AIN2 and 17.4% in AIN1/HPV-infection, with 19 distinct genotypes detected during precancer stages, underscoring the importance of targeting the HPV whole genome and diverse genotypes to improve sensitivity. More broadly, blood-based detection of precancer remains a major challenge for liquid biopsy (*49*). For example, in colorectal cancer screening, the ctDNA-based Shield assay demonstrated only 13.2% sensitivity for detecting advanced precancerous polyps (*8*). Here we leveraged the unique presence of viral cfDNA in HPV-associated cancers and employed a targeted viral whole-genome sequencing strategy, demonstrating meaningful blood-based detection of HPV precancers.

The multi-feature annotation capability of HPV-DeepSeek liquid biopsy further enabled systematic characterization of HPV cfDNA across the HPV infection-precancer-cancer continuum, uncovering stage-informative molecular features while offering insights into the biological processes underlying HPV carcinogenesis. First, we observed a significant increase in HPV cfDNA levels from high-grade precancer to invasive cancer, with median HPV read counts rising from 104 in AIN3/AIN2 to 29,163 in ASCC, and median HPV genome coverage from 48% to 100%. The results align with the exponential growth of tumor burden during progression from precancer to cancer. Second, genomic hallmarks of HPV cancer, including HPV integration events, 3q amplification, and *PIK3CA* mutations, were detectable in blood only among cancer cases. However, the extremely low tumor fraction in blood during precancer stages may have limited the detection of these hallmarks even if they were biologically present. Notably, one AIN3 sample showed HPV cfDNA level close to the ASCC group (13,660 HPV read counts and 100% HPV genome coverage), yet those genomic hallmarks were still not detected. Third, the precancer stages exhibited highly diverse HPV genotypes, whereas cancer cases were predominantly HPV16, consistent with tissue-based studies reporting that 85% of anal cancers are HPV16 while precancer lesions show higher prevalence of other genotypes (*50*). Notably, liquid biopsies are recognized to overcome the sampling bias associated with tissue biopsies, offering a more comprehensive representation of tumor mutational heterogeneity in mutation-based ctDNA assays (*51*, *52*). In our prospective cohort, two samples harbored 7 HPV genotypes in the blood that were distributed across multiple distinct lesion sites, highlighting the same advantage of liquid biopsy in recapitulating the heterogenous viral landscape across lesion sites. This advantage is particularly pronounced in anal cancer and precancer screening, where the complex anatomy of the anal canal and the multifocal nature of HPV diseases pose challenges for comprehensive tissue biopsy (*19*).

Leveraging the lesion-specific origin of HPV cfDNA, we further performed fragmentomics analysis to identify additional features for stage stratification. Our analysis revealed stage-associated remodeling of HPV cfDNA fragmentation patterns across the infection–precancer–cancer spectrum. Compared with precancer stages, ASCC exhibited increasingly defined nucleosomal periodicity, including more pronounced mono- and di-nucleosomal peaks, particularly among integration-positive cases. The observed enrichment of longer, di-nucleosomal fragments in HPV cancer may reflect greater contribution of integrated HPV genomes embedded within host chromatin, whereas the shorter and more heterogeneous fragment profiles observed in earlier stages may be influenced by episomal viral DNA with less defined nucleosomal organization. To enable sample-level annotation of fragmentomics features for stage stratification, we developed a novel likelihood-based framework. We first performed unsupervised clustering of HPV cfDNA fragmentomics profiles within each disease stage, identifying distinct sub-patterns. For each sample, we then computed stage-specific likelihoods based on the similarity of its fragmentomics profile to these stage-defined clusters. This approach accommodates the inter-sample variability and the low HPV cfDNA fragment counts during the precancer stages, enabling improved grouping of samples by disease stage.

The developed liquid biopsy classifier in our study achieved a mean sensitivity of 73.2% and specificity of 82.7% for detecting AIN2+, stages that require clinical intervention, highlighting its potential to address the urgent unmet need in anal cancer screening. The ANCHOR trial demonstrated the clinical benefit of treating anal high-grade precancer in people living with HIV (PLWH), underscoring the importance of anal cancer screening (*21*). Current guidelines from the International Anal Neoplasia Society recommend screening high-risk populations with anal cytology and/or HPV testing (*22*). However, these procedures are invasive, limiting screening compliance. Moreover, cytology is subjective and shows limited sensitivity for detecting anal high-grade precancer and cancer (22.6% in PLWH), while HPV testing or HPV/cytology co-testing show poor specificity (34.9% and 31.5% in PLWH, respectively) (*23*). Critically, there is a significant gap in the availability of HRA, the diagnostic workup following abnormal screening, with an estimated 1/3 PLWH lacking access to HRA referral (*26*). The HRA shortage is projected to worsen further as anal cancer screening becomes standard of care among high-risk populations, substantially increasing the demand for HRA (*24*). Therefore, a liquid biopsy approach with integrated risk stratification could provide a noninvasive alterative for screening high-risk populations or serve as a triage tool following abnormal cytology or HPV testing results, to prioritize patients at elevated risk while safely monitoring others.

It is recognized that not all HPV high-grade precancers are progressive, but current clinical approaches cannot distinguish lesions that are likely to progress from those that may spontaneously regress. In our study, a subset of AIN3/AIN2 samples were consistently assigned high probabilities by the liquid biopsy classifier, raising the intriguing possibility that these lesions may carry a higher risk of progression, whereas those with lower probabilities may be more likely to regress, particularly given that their HPV cfDNA molecular features clustered towards ASCC and AIN1/HPV-infection profiles, respectively. Because all high-grade precancer patients in this study were treated, this hypothesis requires investigation in future independent cohorts without clinical intervention. Additionally, as high-risk populations undergo routine HPV screening surveillance, longitudinal characterization of blood HPV cfDNA, including changes in cfDNA level and associated molecular features, could be used to track disease progression or regression, as well as differentiating new versus persistent infections, to potentially further improve cancer risk prediction.

Our study has several limitations. First, due to the small sample size, the developed classifier was not evaluated in an independent validation cohort. Future studies should assess assay performance in larger sample sets and further optimize the training and validation of the classifier, to ensure its robustness and generalizability. Second, we only investigated the length patterns in fragmentomics features. Emerging studies suggest that the fragment end motifs harbor further biologically informative signals (*44*, *53*). Future work should evaluate the biomarker potential of HPV cfDNA end motifs across the disease stages. Third, in this study we used anal cancer and precancer as our model systems. Future studies should be conducted in additional HPV cancer types to characterize performance metrics.

In summary, our findings demonstrate that blood-based screening and risk stratification of HPV-associated precancer and cancer are feasible, with the potential to improve the accuracy, efficiency, and accessibility of current screening paradigms.

## MATERIALS AND METHODS

### Human subjects

All participants provided written informed consent to protocols approved by the institutional review boards at Dana Farber/Harvard Cancer Center and Mass General Brigham. This study was conducted in compliance with the US Common Rule. Samples from three cohorts of participants were used.

### Biobank cohort

The Mass General Brigham Biobank has prospectively collected blood samples and data from 140,000 patients presenting for care in the healthcare system in Boston, Massachusetts. Biobank clinical data was queried by biobank staff to identify patients who (1) were diagnosed with HPV-associated anal squamous cell carcinoma more than 1 year after blood sample collection; (2) had at least 1 mL of plasma stored; (3) had no history of HPV-associated disease at the time of blood sample collection. A total of 6 patients were identified. Medical records were reviewed to extract clinicodemographic data. Two patients had tumor tissue FFPE samples available at the time of cancer diagnosis which were retrieved for analysis.

### Prospective high-risk screening cohort

We prospectively collected blood samples from patients undergoing anal cancer screening at the Anal Dysplasia Clinic of Mass General Hospital (ClinicalTrials.gov number: NCT06971276). The high-risk screening cohort comprised individuals with past history of HPV infection or treated precancerous lesions who were undergoing HRA. Blood samples were collected at the time of clinical visit and subsequently classified into different disease stages based on standard of care diagnostic results. Specifically, samples from patients negative for clinical HPV testing were classified as anal HPV-negative (N=14). Samples from patients positive for clinical HPV testing were classified into ASCC (N=10), AIN3 (N=20), AIN2 (N=20), AIN1 (N=20), and anal HPV infection without dysplasia (N=20) based on tissue biopsy diagnosis.

### Prospective general population controls

The general population control cohort consisted of 60 patients with no known history of HPV-associated diseases presenting for non-cancer care at Massachusetts Eye and Ear.

### Blood and tissue FFPE sample processing

Blood samples were processed as previously described (*54*). Briefly, 10 to 20 mL of blood collected in DNA BCT tubes (Streck) was double spun at room temperature for 10 minutes at 1,600 g and 3,000 g for plasma isolation. Isolated plasma was kept frozen at -80 °C until use. cfDNA was extracted from 4 mL plasma using the QIAamp Circulating Nucleic Acid kit (Qiagen). For tissue, the lesion site within the FFPE slide was first identified by pathologists. DNA of the lesion site was then extracted using the Quick-DNA FFPE Miniprep kit (Zymo Research) following the manufacturer’s protocol. Extracted DNA was sheared to targeted length of 200 bp via sonication.

### HPV-DeepSeek workflow

The assay was conducted the same as previously described (*16*). Briefly, 20 ng total DNA was used for library preparation following the KAPA HyperCap cfDNA workflow with custom modifications. The main steps were: (1) end repair and A-tailing; (2) KAPA Universal UMI adapter ligation; (3) PCR amplification with KAPA UDI primer mixes; (4) preparation of four-plex library pools for hybrid capture; (5) hybrid-capture enrichment with customized KAPA HyperCap Target Enrichment Probes designed against the whole genomes of 42 HPV genotypes and selected human regions; (6) PCR amplification of the enriched libraries; (7) pooling and normalization of libraries for sequencing. Paired-end sequencing was conducted in Illumina NovaSeq X with roughly 15 million reads per sample.

A custom bioinformatics pipeline was used to process and analyze the sequencing data as previously described with minor modifications. Briefly, sequencing reads first underwent adapter trimming and quality filtering with fastp (version 0.22.0), followed by UMI deduplication with fgbio (version 2.1.0), and aligned to a custom reference genome encompassing hg38 and 64 HPV reference genomes using bwa-mem (version 0.7.17). HPV genotype was determined by the HPV reference genome that the sequencing reads aligned to. The number of unique UMI-deduplicated HPV reads was used as the HPV cfDNA read count output, and the fraction of HPV genome detected was used as the HPV genome coverage output. *PIK3CA* mutations were detected from UMI-deduplicated alignment files. Somatic variants were called using GATK Mutect2 (v4.2.6.1) in tumor-only mode restricted to the *PIK3CA* locus. Variants were annotated with SnpEff (GRCh38.78) and cross-referenced against COSMIC (v100, GRCh38). Clinically relevant *PIK3CA* hotspot mutations were identified by matching annotated calls to a curated COSMIC hotspot list, and variant allele fraction (VAF) was calculated from reference and alternate read counts (VAF = ALT/(REF+ALT)).

The assay threshold for HPV cfDNA was established previously through analytical dilutional experiments, with 10 HPV cfDNA read counts and 10% HPV genome coverage defined as the cutoff threshold for calling HPV cfDNA positive (*16*).

### HPV integration and rearrangement detection

HPV integration and rearrangements were detected from UMI-deduplicated alignment files following the custom bioinformatics pipeline HPV-SIGNAL as described previously (*34*). Briefly, reads were screened for chimeric split reads (supplementary alignments; SA tag) and supporting soft clips (MAPQ >=20). Breakpoints were computed from CIGAR strings, normalized, and filtered to remove HPV reference-end artefacts and self-breakpoints, then clustered by anchoring HPV coordinates and merging partner positions within 10 bp. Integration sites (HPV-human) and intra-viral junctions (HPV-HPV) were reported from clusters supported by at least 6 chimeric reads.

### Long-read WGS sequencing on tissue FFPE samples

100 ng DNA extracted from FFPE tissue samples were used for PacBio long-read sequencing. Libraries were prepared using a combination of PacBio Ampli-Fi and Kinnex workflow to amplify and concatenate FFPE DNA. Specifically, 100 ng input DNA first underwent end-repair and A-tailing, followed by ligation with custom Kinnex adapters. Adapter-ligated DNA were amplified using KOD Xtreme Hot Start DNA polymerase (Millipore Sigma) followed by SMRTbell cleanup. For Kinnex PCR, 25 ng of adapter-ligated DNA was used per reaction with 16-fold concatenation (PacBio). PCR products were pooled and purified prior to array formation and nuclease treatment. Final libraries were assessed for fragment size distribution using the Femto Pulse system and concentration using the Qubit 1x dsDNA HS assay kit. Libraries were subsequently polymerase-bound using the Revio SPRQ polymerase kit and sequenced on the PacBio Revio Platform, with one SMRT Cell per library.

### PCR validation of the HPV integration and rearrangement detection

We designed qPCR assays targeting two selected breakpoints in sample 001, including one HPV-human breakpoint and one HPV-HPV breakpoint. The qPCR assay was performed on DNA extracted from paired plasma and FFPE tissue samples using the Power SYBR™ Green PCR Master Mix (Applied Biosystems) on a QuantStudio™ 6 Flex Real-Time PCR System (Applied Biosystems). Plasma cfDNA from another HPV-associated oropharyngeal cancer patient without the targeted breakpoints was used the negative control. The primer sequences are as following.

Breakpoint 1, forward primer: TGGATGACACAGAAAATGCTAGT

Breakpoint 1, reverse primer: TGGCAAGAAAATGAGGAGTGC

Breakpoint 2, forward primer: ATAAAGTTGGGTGGCCGATG

Breakpoint 2, reverse primer: AGACAGCGGGTATGGCAATA

*GAPDH*, forward primer: ACCACAGTCCATGCCATCAC

*GAPDH*, reverse primer: TCCACCACCCTGTTGCTGTA

### Copy number alteration detection using ichorCNA

For each sample, an off-target bam file was generated by excluding reads mapping to the targeted hybrid-capture regions and other recurrently co-captured loci. We then performed ichorCNA with parameters optimized for low tumor fractions, including setting initial ploidy to diploid, excluding subclonal copy number events, training and analyzing autosomes only, and setting the initial non-tumor fractions as c(0.8, 0.9, 0.95, 0.995, 0.999) (*38*). 3q amplification was called when ichorCNA reported amplification or high-level amplification in chromosome 3 and the amplified segment encompassed the 3q arm but did not span the entire chromosome.

### Fragmentomics analysis

For fragmentomics analysis, UMIs were extracted using a modified fgbio pipeline. Although the standard configuration assumes a fixed 3M3S+T 3M3S+T read structure, our libraries contained variable-length UMIs (4 or 6 bases) followed by a terminal thymine (T) adaptor. Misparsing of this adaptor with the standard configuration resulted in incorrect fragment length profiles. To address this issue, we applied four alternative UMI extraction configurations (4M+T 6M+T, 6M+T 6M+T, 4M+T 4M+T, and 6M+T 4M+T) and merged correctly parsed reads prior to consensus generation. This approach reduced read loss during UMI collapsing, eliminated adaptor-related fragment length artifacts, and produced read counts consistent with the standard HPV-DeepSeek workflow.

From the UMI-processed BAM files, fragments mapping to the HPV genome and the human genome were analyzed separately. For each fragment, the genomic start and end coordinates, total fragment length, and strand orientation were extracted using a custom Python script implemented with the pysam library for downstream fragmentomics analyses. To estimate the mono-nucleosomal peak position, fragment length distributions were smoothed using a Gaussian filter (σ = 3 bp), and the peak position was defined as the mode of the mean smoothed curve within the 120-200 bp range.

For statistical testing under sparse fragment counts and small group sizes, we aggregated fragment counts across samples within each group to construct a stable group-level fragment-length spectrum. Fisher’s exact tests (bin vs. non-bin) were then performed to assess bin-level enrichment. This approach was chosen because many precancer samples contained very few HPV fragments, resulting in highly discretized and stochastic bin proportions at the individual-sample level. For consistency in statistical testing, and to accommodate small-group comparisons (n = 6 vs. n = 4), the same aggregated-count strategy was applied to the integration-status analysis.

For the three disease stage groups (AIN1/HPV Infection, AIN3/AIN2, and ASCC), samples were independently clustered using hierarchical clustering with Euclidean distance and Ward linkage, yielding k=4, 3, and 2 clusters, respectively. Within each cluster, HPV fragment counts across samples were aggregated and normalized by the total fragment counts to produce average cluster probability distributions. For each sample, multinomial likelihood scores were computed against each of the 9 clusters by comparing the sample’s HPV fragment length distribution to the reference cluster probability distributions. Finally, the posterior probabilities were summed across all samples within each disease stage group to obtain total cluster likelihoods per group.

### Training and validation of the integrated classifier

We used elastic net-regularized logistic regression model to develop a binary classifier for distinguishing intervention groups (ASCC, AIN3, and AIN2) from the non-intervention groups (AIN1, HPV-infection, and HPV-negative) within the high-risk screening cohort. The model features included HPV read counts, HPV genome coverage, presence of high-risk HPV genotype, presence of HPV integration or rearrangement events, presence of 3q amplification, presence of *PIK3CA* mutations, and fragmentomics likelihoods to ASCC, AIN3/AIN2, and AIN1/HPV-infection. Model training and evaluation was conducted in the R glmnet package within the caret framework, using nested 5-fold cross-validation repeated 5 times. Model hyperparameters were selected based on AUC. The ROC curve, AUC, and sensitivity and specificity at the Youden index threshold across all splits (25 in total) were used to generate the mean performance and the 95% confidence intervals of the model.

For each sample, predicted probabilities from each outer fold were summarized across the 5 repeats. To mitigate variability across folds, probabilities in each outer fold were normalized relative to the corresponding inner-fold sample probabilities using the empirical cumulative distribution function. In addition, sample classification (positive versus negative) in each outer fold was determined using the Youden index threshold derived from the corresponding inner fold.

### Statistics

One-sided Wilcoxon rank-sum tests were used to assess the statistical significance of increases in HPV cfDNA read counts and HPV genome coverage with progression from AIN1/HPV infection to AIN2/3 and ASCC (**Figure 2e**). Fisher’s exact tests were used to evaluate differences in detection rates of HPV structural variations, 3q amplification, and *PIK3CA* mutations between ASCC and non-ASCC cases (**Figures 3b, 3h, and 3j**). Fisher’s exact tests with Benjamini–Hochberg correction for multiple testing were applied to fragmentomics enrichment comparisons (**Figures 4c and 4e**).

## Data Availability

Raw data (FASTQ files) and processed data (BAM files) generated from the study will be deposited to dbGaP upon manuscript publication.

## Author contributions

DLF conceived and acquired funds for the study. DLF, BTD, and DCG supervised the study. QW, DD, YA, and SH conducted wet-lab experiments. QW, SE, SSL, and ER conducted bioinformatics analysis. QW, SE, and SSL led the formal analysis. GL, ETE, SS, MGD, and BTD procured clinical samples. HD and JL procured the long-read tissue WGS data. VAA provided resources and supervision on the methodology. QW, SSL, DCG, and DLF wrote the first draft of the manuscript. All authors participated in revision of the manuscript. Authorship order among co-first authors (QW, SE, and SSL) was determined based on the relative contribution to conceptualization, data curation and analysis, and manuscript writing.

## Data, code, and materials availability

Raw data (FASTQ files) and processed data (BAM files) generated from the study will be deposited to dbGaP upon manuscript acceptance. Source data used for the figures and supplementary figures are provided in the auxiliary data file. Tools used for data analysis are publicly available from the indicated references.

