## Supplementary Information for "Liquid Biopsy of HPV Cell-Free DNA Enables Blood-Based Early Detection and Molecular Stratification of HPV-Associated Cancer and Precancer Stages"

**Supplementary Table 1. Demographics of the biobank cohort.**

| | Patient No. (%) n=6 |
|---|---|
| **Age at ASCC diagnosis, median (range)** | 60 (52 – 82) |
| **Sex** | |
| - Male | 0 (0%) |
| - Female | 6 (100%) |
| **Clinical T stage** | |
| - 0 | 0 (0%) |
| - 1 | 0 (0%) |
| - 2 | 2 (33.3%) |
| - 3 | 2 (33.3%) |
| - Details not available | 2 (33.3%) |
| **Clinical N stage** | |
| - 0 | 3 (50%) |
| - 1 | 1 (16.7%) |
| - Details not available | 2 (33.3%) |
| **Clinical M stage** | |
| - 0 | 4 (66.7%) |
| - Details not available | 2 (33.3%) |
| **Plasma collection lead time, median (range)** | 4.5 (2.8 – 8.6) |

**Supplementary Table 2. Demographics of the prospective high-risk screening cohort.**

| | Patient No. (%) n=104 |
|---|---|
| **Age at diagnosis, median (range)** | 53.5 (28 – 79) |
| **Sex** | |
| - Male | 90 (86.5%) |
| - Female | 14 (13.5%) |
| **HIV status** | |
| - Positive | 67 (64.4%) |
| - Negative | 37 (35.6%) |
| **Diagnosis** | |
| - ASCC | 10 (9.6%) |
| - AIN3 | 20 (19.2%) |
| - AIN2 | 20 (19.2%) |
| - AIN1 | 20 (19.2%) |
| - Anal HPV-infection | 20 (19.2%) |
| - Anal HPV-negative | 14 (13.5%) |
| **ASCC, clinical T stage (n=10 total)** | |
| - 1 | 1 (10%) |
| - 2 | 5 (50%) |
| - 3 | 1 (10%) |
| - 4 | 3 (30%) |
| **ASCC, clinical N stage (n=10 total)** | |
| - 0 | 4 (40%) |
| - 1 | 4 (40%) |
| - Details not available | 2 (20%) |
| **ASCC, clinical M stage (n=10 total)** | |
| - 0 | 10 (100%) |

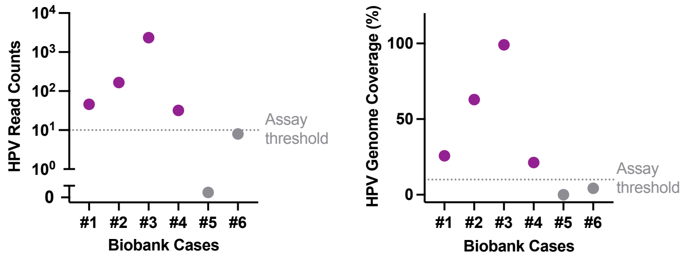


**Supplementary Figure 1. HPV cfDNA level in biobank plasma samples**.

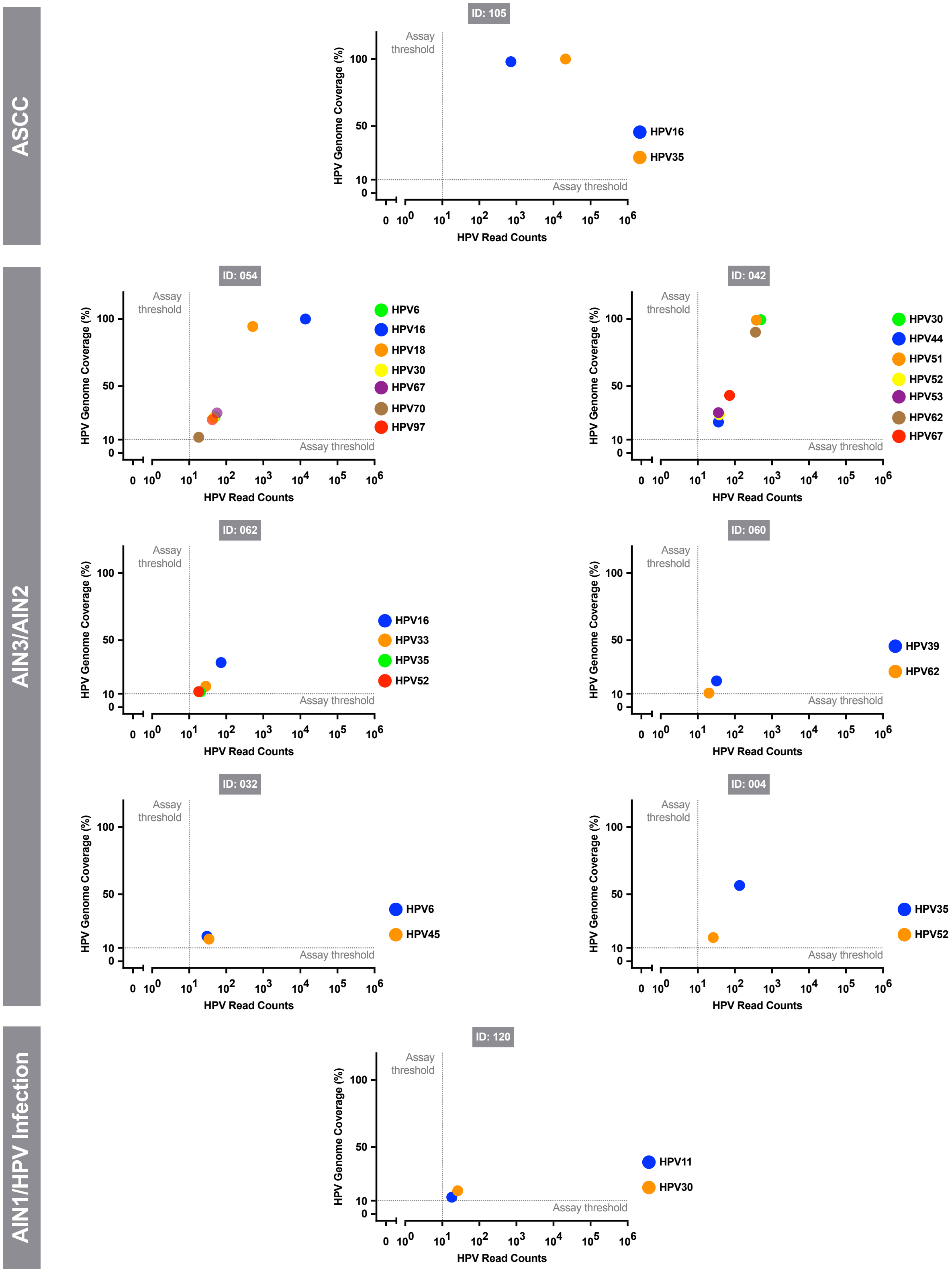


**Supplementary Figure 2. HPV cfDNA level of each genotype in the multi-genotype cases.**

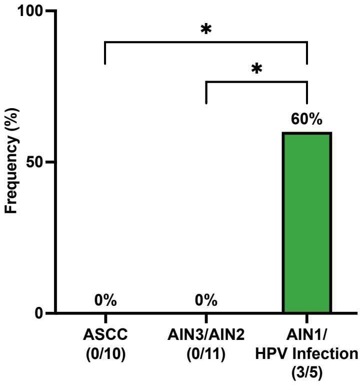


**Supplementary Figure 3. Frequency of cases harboring only low-risk HPV genotypes across the different disease groups.** Statistical significance was determined using Fisher's exact test (**p* < 0.05).

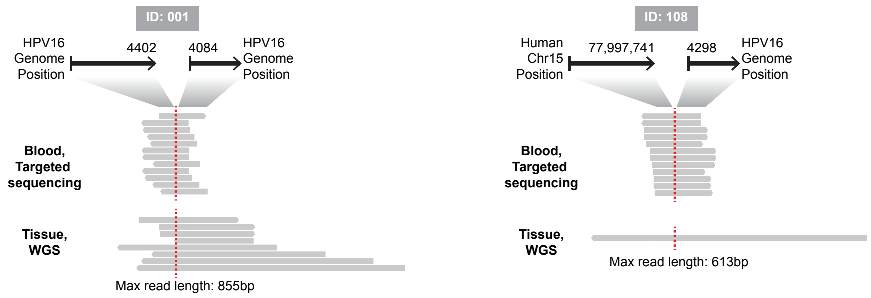


**Supplementary Figure 4. Tissue WGS validation of breakpoints identified in paired plasma samples.**

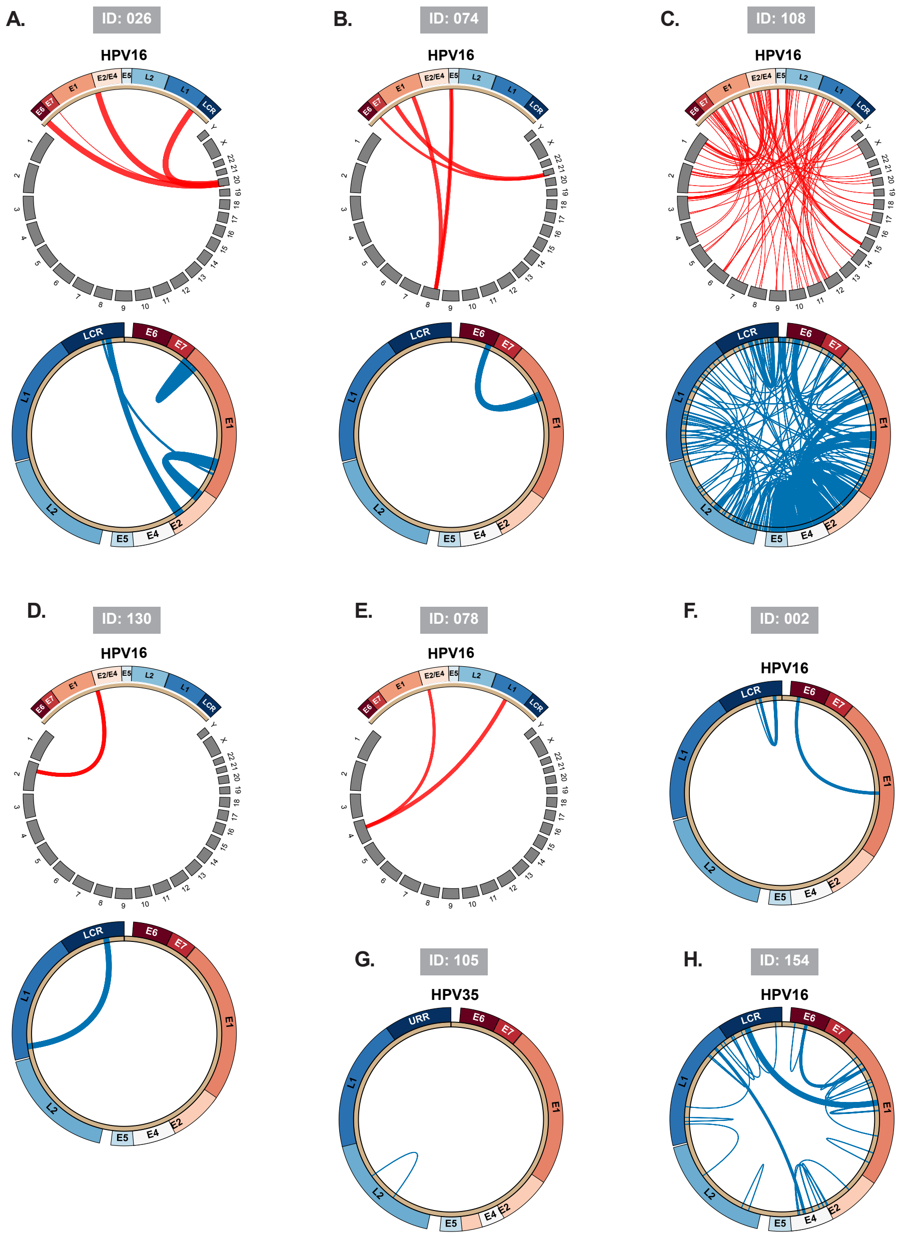


**Supplementary Figure 5. HPV-human integration and HPV-HPV rearrangement details in all detected cases.** The HPV genome is shown as a sector annotated with viral genes, with human chromosomes shown as gray sectors. Red links indicate HPV–human breakpoints and blue links indicate HPV-HPV non-circular junctions. Link width scales with the number of supporting reads.

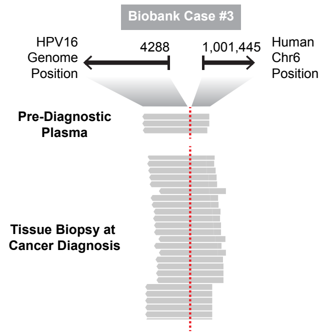


**Supplementary Figure 6. HPV-human integration events found in the pre-diagnostic plasma that was validated in the tissue biopsy sample at cancer diagnosis in the biobank cohort.**

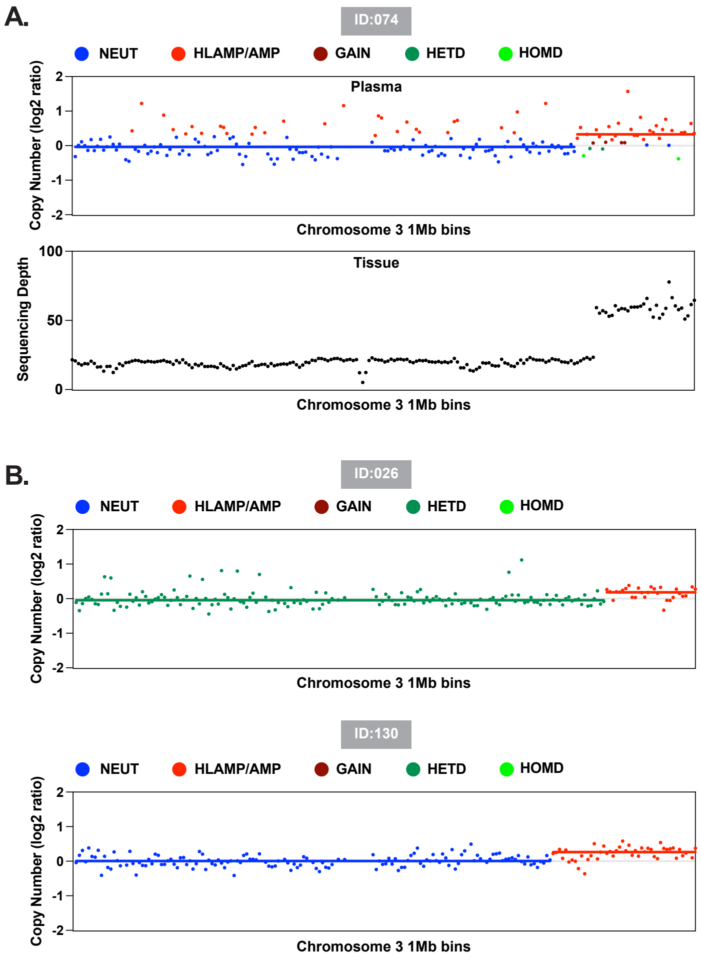


**Supplementary Figure 7. Details of cases with 3q amplification detected in plasma.** (A) One sample with paired tissue WGS showing concordant 3q amplification. (B) Two additional samples with 3q amplification detected in plasma. HOMD, HETD, NEUT, GAIN, AMP, HLAMP refer to states called by ichorCNA and defined as the following: HOMD, homozygous deletions; HETD, hemizygous deletions; NEUT, copy neutral; GAIN, copy gain; AMP, amplification; HLAMP, high-level amplification.

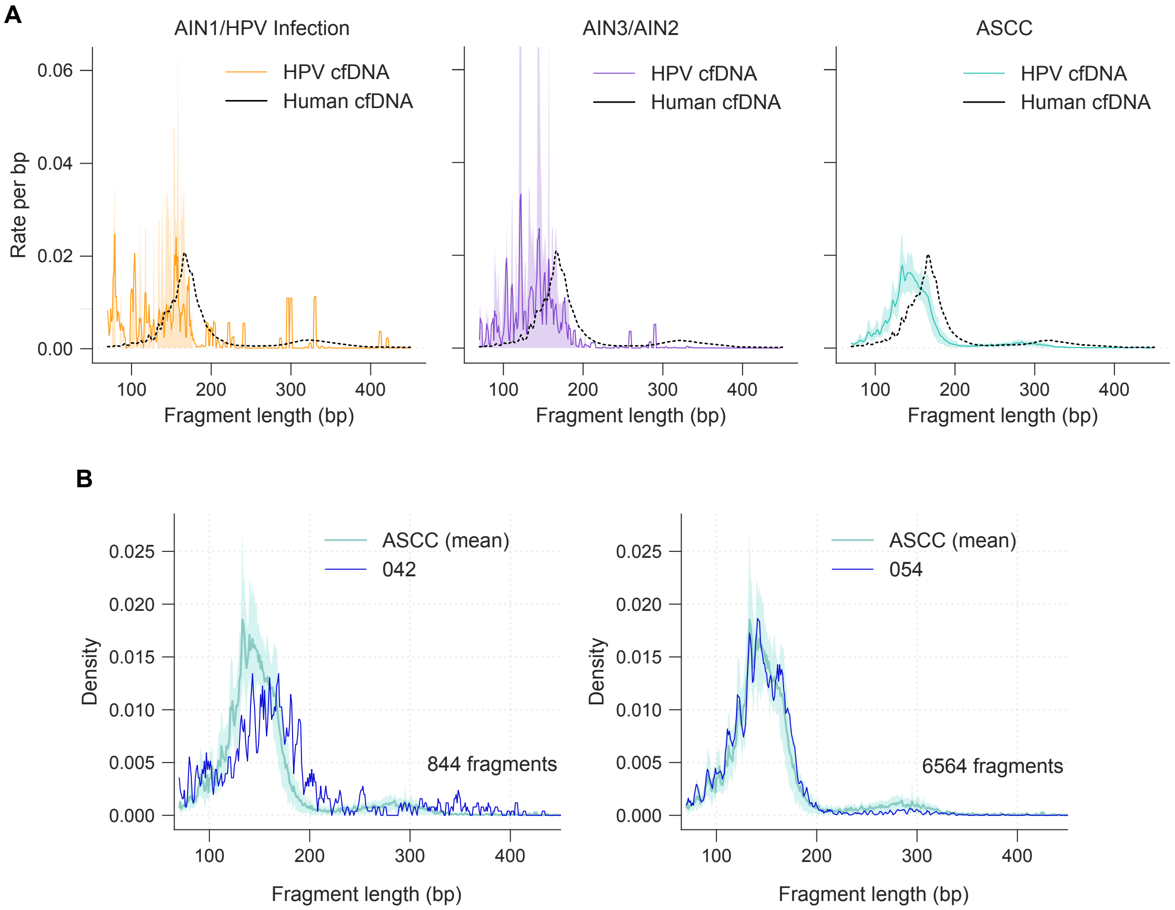


**Supplementary Figure 8. HPV cfDNA fragment-length profiles. (A)** Group-level fragment-length profiles for anal AIN1/HPV infection, AIN3/AIN2, and ASCC. Fragment length distributions are shown as rate per base pair for HPV-derived cfDNA (colored) and human cfDNA (black). For visualization, HPV-derived cfDNA curves were smoothed using a 3-bp centered rolling mean. **(B)** Fragment-length profiles of two AIN2/AIN3 outlier samples (042 and 054) with unusually high HPV fragment counts, which were excluded from the analyses in Fig. 4. Individual HPV cfDNA rate profiles (smoothed using a 3-bp centered rolling mean) are shown overlaid on the ASCC group mean HPV cfDNA rate (teal) for reference.

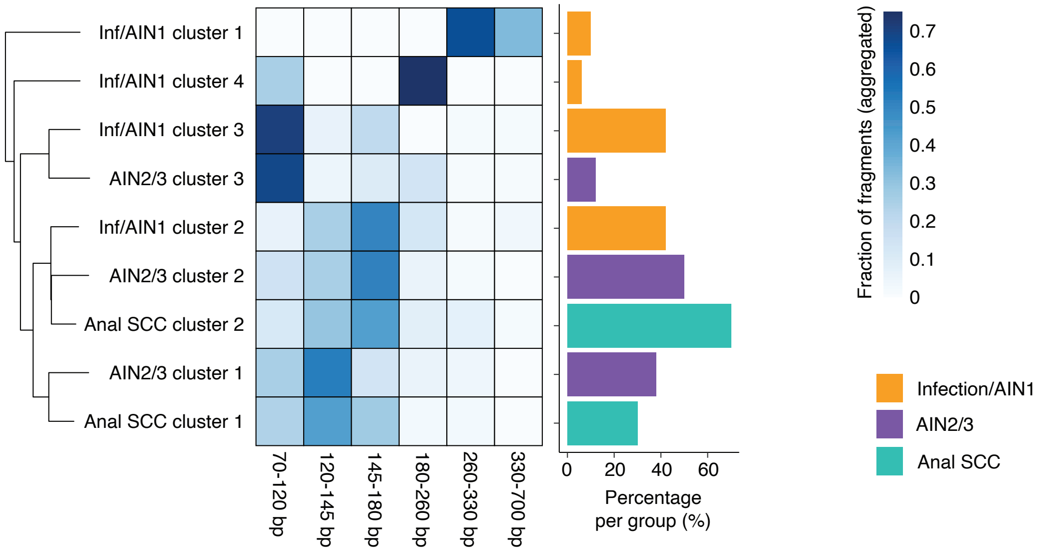


**Supplementary Figure 9. Fragmentomics clusters based on length distribution patterns.** For the stage groups (Infection/AIN1, AIN2/AIN3, and Anal SCC), we clustered samples, finding k = 4, 3, and 2 clusters, respectively. Then the fragments in samples are aggregated for each cluster and are normalized by the total fragment counts to calculate average cluster probability distributions shown in the heatmap (left panel). The number of samples per cluster is shown in the bar plot next to the probability distributions.

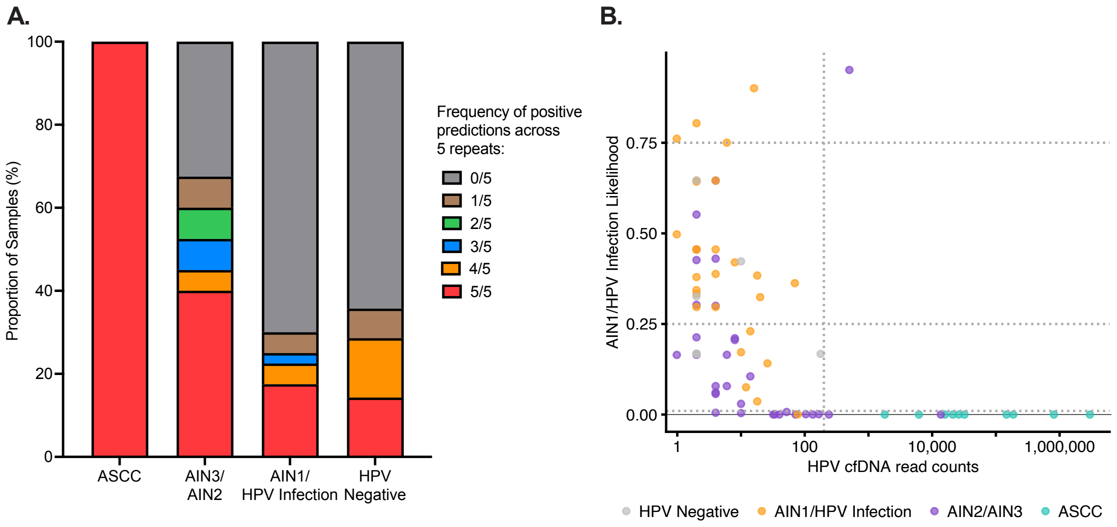


**Supplementary Figure 10. Potential of the classifier in annotating underlying cancer risk. (A)** Proportion of samples with different frequencies of being predicted positives across the 5 repeats within each disease stage. **(B)** Scatter plot showing the distribution of samples by fragmentomics AIN1/HPV-infection likelihoods and HPV cfDNA fragment counts. 8 regions were defined using thresholds of 0.01, 0.25, and 0.75 for the fragmentomics AIN1/HPV-infection likelihoods and a threshold of 200 for HPV cfDNA fragment counts.
